# Cardiac resynchronization therapy among adults with a systemic right ventricle: a multicenter experience

**DOI:** 10.1101/2025.03.06.25323532

**Authors:** Flavia Fusco, Giancarlo Scognamiglio, Mikael Dellborg, Payam Dehghani, Susan M. Jameson, W. Aaron Kay, Jonathan Cramer, Isabelle Vonder Muhll, Eric V. Krieger, Fred Rodriguez, Luke J. Burchill, Jeremy Nicolarsen, Joseph Kay, Robert M. Kauling, Sangeeta Shah, Anthony Magalski, Joshua Wong, David S. Celermajer, David Baker, Jolien Roos-Hesselink, Salil Ginde, Jamil Aboulhosn, Marissa Kuo, Christopher DeZorzi, Paul Khairy, Carla P. Rodriguez-Monserrate, Shelby Kutty, William M. Wilson, Adam M. Lubert, Jasmine Grewal, Frank Han, Timothy Cotts, Stephen Pylypchuk, Tripti Gupta, Petra Antonova, Clare O’Donnell, Anitha S. John, Pastora Gallego, Alexandra van Dissel, Alexander R. Opotowsky, Elizabeth Yeung, Craig S. Broberg, Berardo Sarubbi

## Abstract

**Background:** Cardiac Resynchronization Therapy(CRT) is a key treatment for heart failure (HF) in acquired heart disease, but its benefits in adults with congenital heart disease and a systemic right ventricle (sRV) remain unclear. This study aimed to assess whether CRT improves outcomes in patients with sRV.

**Methods:** This analysis was part of an international, retrospective study of data from 33 centers including patients >18years with transposition of the great arteries (TGA) following atrial switch operation and congenitally corrected TGA (ccTGA). The primary endpoint was overall survival. The secondary endpoint was a composite of death, hospitalization for HF, heart transplant, mechanical support, ventricular tachycardia/ICD therapies.

**Results:** We identified 105 out of 1,721 patients(3.5%) who underwent CRT. Median follow-up after CRT implant was 4.6 [1.6-8] years. There was no overall QRS improvement (157±37 Vs 153±31 ms; p=0.2), which was limited to those with previous pacing (167±35 Vs 154± 28 ms; p=0.002). Following CRT, there was no significant change in B-type natriuretic peptide values, peakVO_2_, tricuspid regurgitation severity by echocardiography. CRT complications occurred in 10 (9.5%), though there were usually minor. Patients with CRT were propensity-matched to controls according to age, sex, morphology (ccTGA/TGA), presence of complex disease, previous HF admission and sRV dysfunction at baseline. At univariable analysis, CRT (OR 4.02-95% CI:1.48-10.89; p=0.006), older age, and moderate to severe sRV dysfunction at baseline were predictive of death. By multivariable analysis, CRT (OR 2.1-95% CI:1.2-3.8; p=0.008), age (OR 1.03-95% CI:1.01-1.06; p=0.001) and moderate to severe sRV dysfunction (OR 2.5-95% CI:1.3-4.4; p=0.002) were independently associated with poorer outcome. After matching using only patients with subpulmonary ventricle pacing as controls, overall survival was still worse in CRT group (p=0.0057).

**Conclusion:** In this retrospective study in the largest population thus far described with a sRV, CRT implant was not associated to improved survival, even after controlling for key confounders. Further studies with randomization are required to improve the selection of candidates with a sRV most likely to derive benefit from CRT, elucidate the optimal timing, and assess alternative strategies.

## 1. Introduction

Cardiac Resynchronization Therapy (CRT) improves heart failure (HF) outcomes for a subset of patients with left ventricular dysfunction due to acquired heart disease. Prevalence of HF in the adult congenital heart disease (ACHD) population, particularly among those with a systemic right ventricle (sRV) is increasing (1–2). As a result, there is a growing need to establish the optimal treatment strategy (3). Recently, promising results have been demonstrated by novel HF medications in this population (4–6). However, there remains uncertainty regarding efficacy, safety, and patient selection for HF electrical therapy in this unique population, especially considering the challenges of CRT implantation due to unconventional anatomy and associated lesions. Published experience is limited by small sample sizes, heterogeneous patient populations, and variable follow-up durations. However, studies to date have not demonstrated a survival benefit from CRT in sRV (7). We aimed to address these knowledge gaps by conducting an international multicenter retrospective analysis of CRT outcomes in patients with a sRV.

## 2. Methods

### 2.1. Data collection

The current study was conducted as part of an international, multicenter, retrospective cohort study executed by the Alliance for Adult Research in Congenital Cardiology (AARCC) on outcomes of adults with a sRV. Collaborators from ACHD centres globally participated after approval by local ethics oversight. Informed consent was waived given the retrospective nature of the study design.

Variables and data collection specifics have been previously published (1). Eligible adult patients (age ≥18 years) included those with congenitally corrected TGA or d-TGA who had undergone previous atrial switch procedure (Mustard or Senning). Patients were required to have been seen at least twice by an ACHD specialist and followed for at least one year. We excluded patients who no longer had a systemic RV (i.e. arterial switch, Rastelli procedure, etc). Complex anatomy designation included coexisting valvular or subvalvular pulmonary stenosis, double outlet RV, or laterality defects (i.e. dextrocardia or situs abnormalities). The first outpatient ACHD encounter since January 1, 2002 was identified as the initial visit, and the last known ACHD visit was considered the follow-up. Similar data were collected for both the initial and follow-up visits. In addition, any major events since the initial visit were recorded, including arrhythmias, new pacemakers, or implantable cardioverter-defibrillators (ICD), or HF admission defined as nonelective hospitalization for HF symptoms with elevated B-type natriuretic peptide (BNP) or need for diuresis.

Specific data were obtained for all patients in whom CRT had been attempted or established. These included indication for CRT, date of attempt, medications at the time of attempt, lead type, and QRS duration before and after successful CRT and any subsequent optimization, as well, lead complications including lead revisions. Data analysis for the present substudy was performed at Monaldi Hospital, Naples, Italy.

### 2.2. Endpoints

The primary endpoint was overall survival. The secondary endpoint was a composite of death, hospitalization for HF, heart transplant (HT), MCS implant, aborted sudden cardiac death, ventricular tachycardia/ICD therapies.

### 2.3. Statistical analysis

Statistical analysis was carried out using R version 4.0.5. Continuous variables were reported as mean±SD or median[IQR], according to data distribution. Patients were divided into 2 groups according to the history of CRT implant: baseline timepoint for survival analysis was set at the time of CRT implant for the CRT group and at study inclusion for those with no CRT. Comparisons between groups were assessed with Student *t-*tests for unpaired samples or Mann-Whitney tests. Categorical variables were presented as frequencies AAQ(percentage of total). Differences in proportions were evaluated with χ^2^ tests. Propensity scores were computed using logistic regression, with presence of CRT as dependent variable and clinical variables as independent variables. The latter included age, sex, anatomy (ccTGA/TGA), complex disease, previous admission for heart failure and sRV systolic function at baseline (assessed by echocardiography and categorized in normal-mildly reduced or moderately-severely reduced systolic function) to perform 1:1 nearest neighbour matching within a caliper of 0.25SDs. Once balance was verified, matched pairs were used as strata in the Cox model. In order to explore the different efficacy of CRT therapy compared to ventricular pacing in sRV, matching was repeated including only patients with ventricular pacing as controls. Univariable Cox proportional hazards regression analysis, with censoring at the time of last follow-up, was used to identify clinical and demographic variables associated with events. Mutivariable models were computed, when feasible. These included all predictors associated with a univariable p-value <0.1, as well as the clinically relevant variables based on substantive knowledge, after which stepwise backward selection was performed with the best model determined by Akaike Information Criteria (AIC). Kaplan–Meier survival curves of matched patients were plotted with censoring the time of last follow-up and time to first events and compared using log-rank tests. In order to compare survival free from events before and after CRT implant, Kaplan-Meier curves were obtained using time of the time of study inclusion as baseline before CRT. P-value<0.05 was considered statistically significant.

## 3. Results

### 3.1. Study population

A total of 1721 patients with a sRV were included from 33 centres worldwide. A CRT implantation procedure was attempted in 114 (7%) patients, 17 (15%) prior to study enrolment and 97 (85%) during follow-up. Baseline patient characteristics are summarized in Table 1. Patients in the CRT group were older and more likely to have ccTGA and complex defects (p<0.0001 for all). CRT patients showed a higher prevalence of atrial fibrillation (p=0.0006) and ventricular pacing (p<0.0001) at baseline and had a higher prevalence of previous ventricular tachycardia (p<0.0001). Accordingly, a higher proportion of CRT patients had a previous ICD (p=0.001) or pacemaker implant (p<0.0001). CRT patients had longer baseline QRS duration (157±38 Vs 113± 27.3 ms; p<0.0001). Data on baseline sRV systolic function estimated by echocardiography were available for 108/114 CRT patients: severely impaired systolic function was present in a higher proportion in CRT patients compared to the group with no CRT (20% Vs 8%; p<0.0001). For those with exercise data reported, CRT patients had lower baseline peak VO_2_ (p<0.0001, N= 52 for the CRT group compared with 684 controls).

**Table 1:**
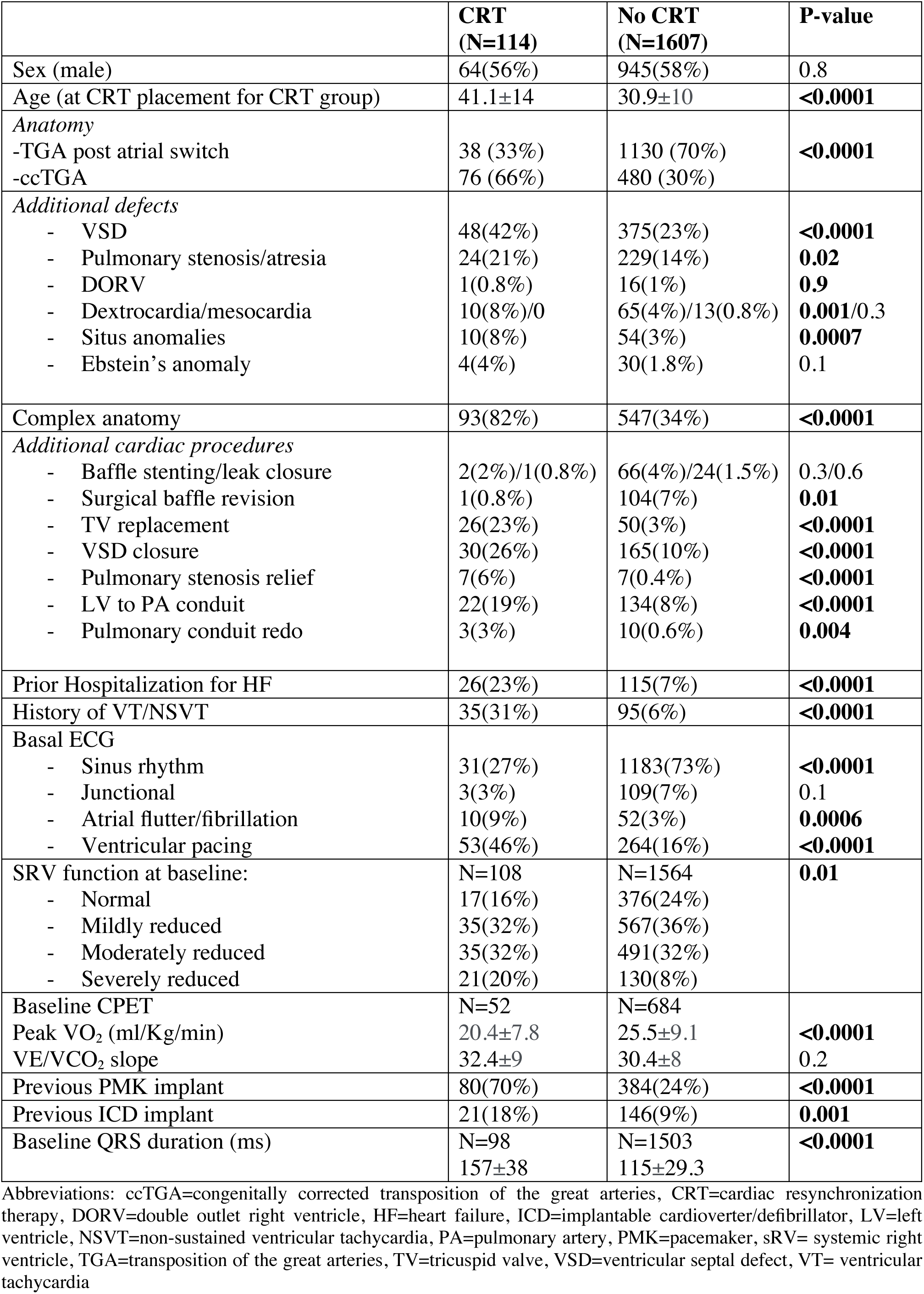
Study population baseline characteristics stratified by candidacy to CRT implant.

### 3.2. CRT implant

In 9 of the 114 patients with attempted CRT (8%), technical reasons precluded successful CRT placement. The remaining 105 individuals had successful CRT implantation. CRT implant was performed after a median of 6.2±4.0 years from the first evaluation. Details on CRT implant procedures are reported in Table 2. In 12 (11%) patients, CRT implant was performed at the time of other cardiac surgery. In the majority of cases (57%), the indication for CRT was worsening HF. Forty-one percent of patients received an upgrading from a single/dual chamber pacemaker after a median of 14.7[6.8-24.6] years from the first implant, leading to a total of 1096.83 person-years spent with subpulmonary ventricular pacing before CRT.

**Table 2.** CRT implant.

|  |  |
| --- | --- |
| Failed CRT placement | 9 (8%) |
| Time from basal evaluation to CRT placement | 6.16±4y |
| Indication to CRT<br>- Pacing needed<br>- Worsening HF<br>- Pacing and HF<br>- Secondary prevention<br>- Unknown | 2(2%)<br>60(57%)<br>31(29%)<br>10(10%)<br>2(2%) |
| Implant type (including attempts)<br>- CRT-P<br>- CRT-D<br>- Upgrade from PMK to CRT-P<br>- Upgrade from ICD to CRT-D<br>- Upgrade from PMK to CRT-D<br>- Unknown | 10(10%)<br>26(25%)<br>35(33%)<br>24(22%)<br>8(8%)<br>2(2%) |
| Time from PMK to CRT upgrading | 14.7[6.8-24.6]y |
| Time from ICD to CRT upgrading | 5.4[2.8-10]y |
| Subsequent upgrade of CRT-P to CRT-D | 1(0.9%) |
| Other procedures performed with CRT implant<br>- TV repair/replacement<br>- Baffle surgical revision<br>- PV/LV-to-PA conduit replacement<br>- MV replacement<br>- Aortic valve replacement<br>- Epicardial PMK lead revision | 12(11%)<br>5<br>1<br>4<br>1<br>3<br>1 |
| Type of CRT lead<br>- Transvenous (CS)<br>- Epicardial or hybrid<br>- Unknown | 36(34%)<br>19(18%)<br>50(47%) |
| Type of capture<br>- Bipolar<br>- Quadripolar<br>- Unknown | 22(21%)<br>10(10%)<br>73(68%) |

Abbreviations: CRT=cardiac resynchronization therapy, CRT-D= cardiac resynchronization therapy – defibrillator, CRT-P= cardiac resynchronization therapy – pacemaker, CS=coronary sinus, HF=heart failure, ICD=implantable cardioverter/defibrillator, LV=left ventricle, MV=mitral valve, PMK=pacemaker, PA=pulmonary artery, PV=pulmonary valve.

### 3.3. Follow-up after CRT placement

Median follow-up after CRT implant was 4.6[1.6-8] years. Complications due to CRT occurred 10 (9.5%), with 8 (8%) requiring lead revision (Table 3). For the entire cohort, QRS duration did not improve after CRT (157±38 Vs 153± 31 ms; p=0.2). For those who were previously paced, the QRS changed from 167±35 Vs 154± 28 ms; p=0.002; for those who were not previously paced, the QRS changed from 133.9±31 Vs 149.7 ± 39 ms; p=0.01. Echo-derived data on sRV systolic function were available both at baseline and follow-up for 100 patients: moderately to severely impaired sRV was present in 51% patients at baseline and in 76% at latest follow-up (p=0.0002, Figure 1). During follow-up, there was no significant change in BNP values, peakVO_2_, tricuspid regurgitation severity (Table 4), though these metrics were not uniformly acquired.

**Figure 1.**
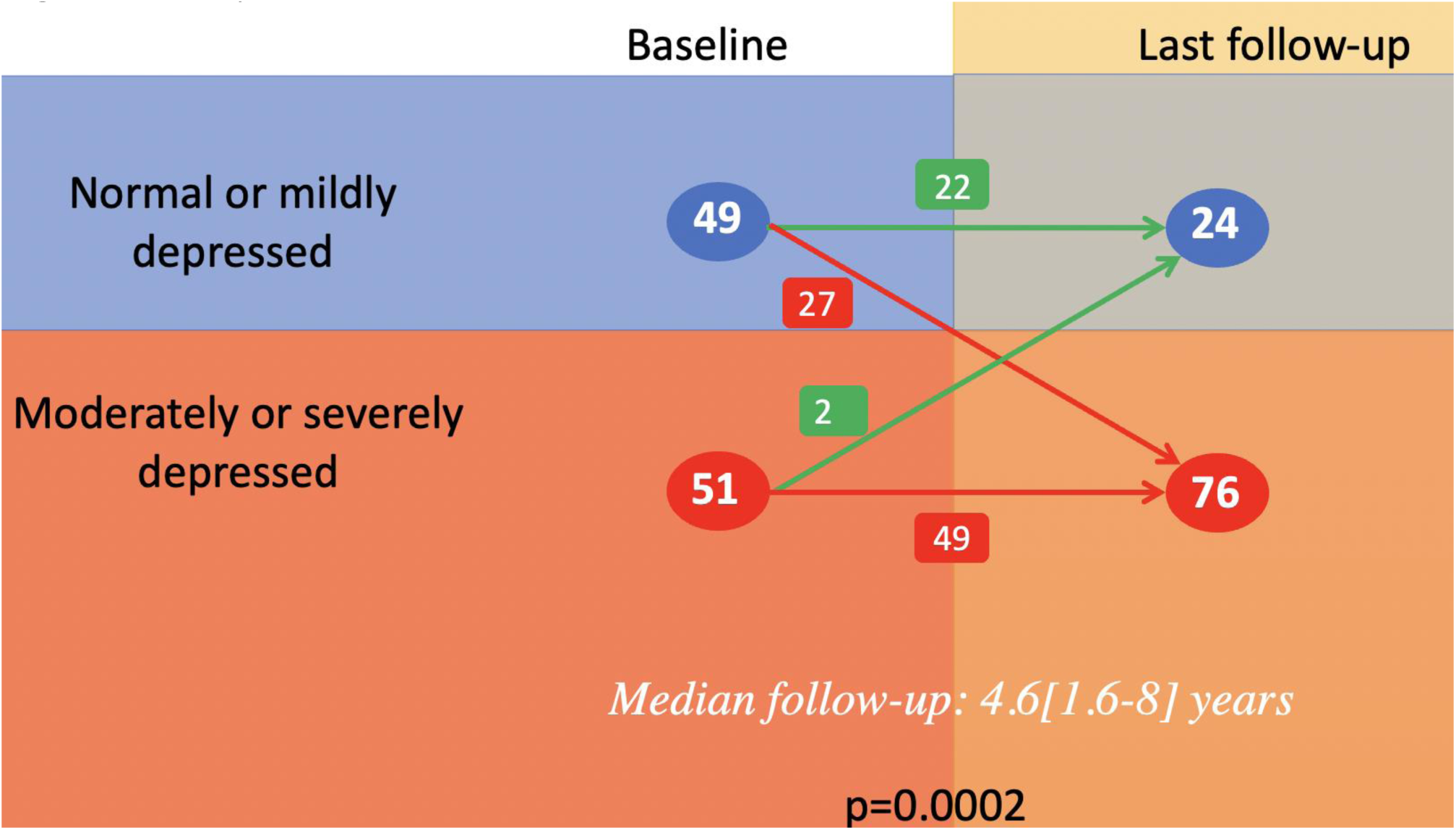
sRV systolic function at baseline and last follow-up in CRT group Data pre and post were available for 100 patients.

**Table 3.** Follow-up after CRT implant.

|  |  |
| --- | --- |
| Follow-up time after CRT | 4.6[1.6-8]y |
| Tricuspid regurgitation (N=103)<br>- None<br>- Mild<br>- Moderate<br>- Severe | 18(17%)<br>39(38%)<br>29(28%)<br>17(17%) |
| SRV systolic function (N=103)<br>- Normal<br>- Mildly reduced<br>- Moderately reduced<br>- Severely reduced | 8(8%)<br>18(17%)<br>45(44%)<br>32(30%) |
| Complications<br>- Site infection<br>- Lead infection and dysfunction<br>- Lead dysfunction<br>- Lead displacement<br>- Diaphragmatic pacing | 10(10%)<br>1(1%)<br>1 (1%)<br>2(2%)<br>4(4%)<br>2(2%) |
| Lead revision | 9(9%)<br>1(1%) for SVC obstruction<br>1(1%) for progressive MR<br>1(1%) during surgery for TV replacement<br>1(1%) extraction and epicardial PMK placement due to infection<br>5(%) Unknown cause |
| Cardiac procedures following CRT (other than revision)<br>- TV replacement<br>- PV replacement<br>- MV replacement<br>- Baffle stenting<br>- Baffle surgical revision | 7(7%)<br>1(1%)<br>2(2%)<br>1(1%)<br>3(3%)<br>2(2%) |

Abbreviations: CRT=cardiac resynchronization therapy, MR=mitral regurgitation, MV=mitral valve, PV=pulmonary valve, PMK=pacemaker, SVC=superior vena cava, SRV=systemic right ventricle, TV=tricuspid valve

**Table 4.** Comparison pre and post CRT implant.

| **Pre Vs Post comparison** |  |  |  |
| --- | --- | --- | --- |
|  | **Baseline** | **Last follow-up** | **p-value** |
| Peak VO<sub>2</sub> (ml/Kg/min) N=34 | 20.5±7.6 | 18.3±6 | 0.2 |
| BNP/upper reference limit N=26 | 3.6[0.7-9.4] | 3.3]1-6.7] | 0.4 |
| QRS duration (ms) N=87 | 157±38 | 153± 32 | 0.2 |
| Moderate to severe TR N=100 | 43 (43%) | 44 (44%) | 0.9 |

Abbreviations: CRT=cardiac resynchronization therapy, BNP=B-type natriuretic peptide; TR=tricuspid regurgitation

### 3.4. Outcome in patients with and without CRT

Events at follow-up are summarized in Table 5: proportion of patients experiencing adverse events was similar in the group of patients with and without CRT. Moreover, no significant differences in events could be demonstrated between CRT patients de novo implant or previous ventricular pacing or in those with an epicardial device compared to transvenous leads (Supplemental table 1), nor in TGA compared to ccTGA group (Supplemental table 2). All CRT patients were matched 1:1 in a propensity score analysis with controls according to above-specified baseline clinical variables. Univariable analysis results are summarized in Table 6: mortality was associated with CRT (OR 4.02 - 95%CI: 1.48-10.89; p=0.006), older age (per +1 year, OR 1.08 - 95%CI: 1.04-1.12; p<0.0001), and moderate to severe sRV dysfunction at baseline (OR 3.6 - 95%CI: 1.2-11; p=0.02). Kaplan-Meier curves showed worse survival in CRT patients (p=0.0032, Figure 2A). Secondary endpoint was significantly associated with CRT, age, and moderate to severe sRV dysfunction on univariable analysis. Multivariable analysis showed that CRT (OR 2.1 - 95%CI: 1.2-3.8; p=0.008, age (OR 1.03 - 95%CI: 1.01-1.06; p=0.001) and moderate to severe sRV dysfunction (OR 2.5 - 95%CI: 1.3-4.4; p=0.002) were independently associated with secondary endpoint. Kaplan-Meier curves showed worse overall event-free survival in CRT patients (p=0.025, Figure 2B). Matching was repeated including baseline QRS duration in addition to all the clinical variables already used: 86 CRT patients were matched to controls and Kaplan-Meier curves depicted similar survival but confirmed worse survival free from events in the CRT group (Supplemental Figure 1). Univariable and multivariable analysis with matching limited to patients with ventricular pacing as controls yielded similar results (Table 7). Kaplan-Meier curves showed worse overall survival and a tendency to significantly worse survival free from events in CRT patients compared with those with subpulmonary pacing (p=0.0057 and 0.06, respectively, Figure 2C-D). To further ascertain whether CRT implant affected the outcome, KM were computed using data collected before CRT for comparison (Supplemental Figure 2): we found no significant differences between pre and post CRT.

**Figure 2.**
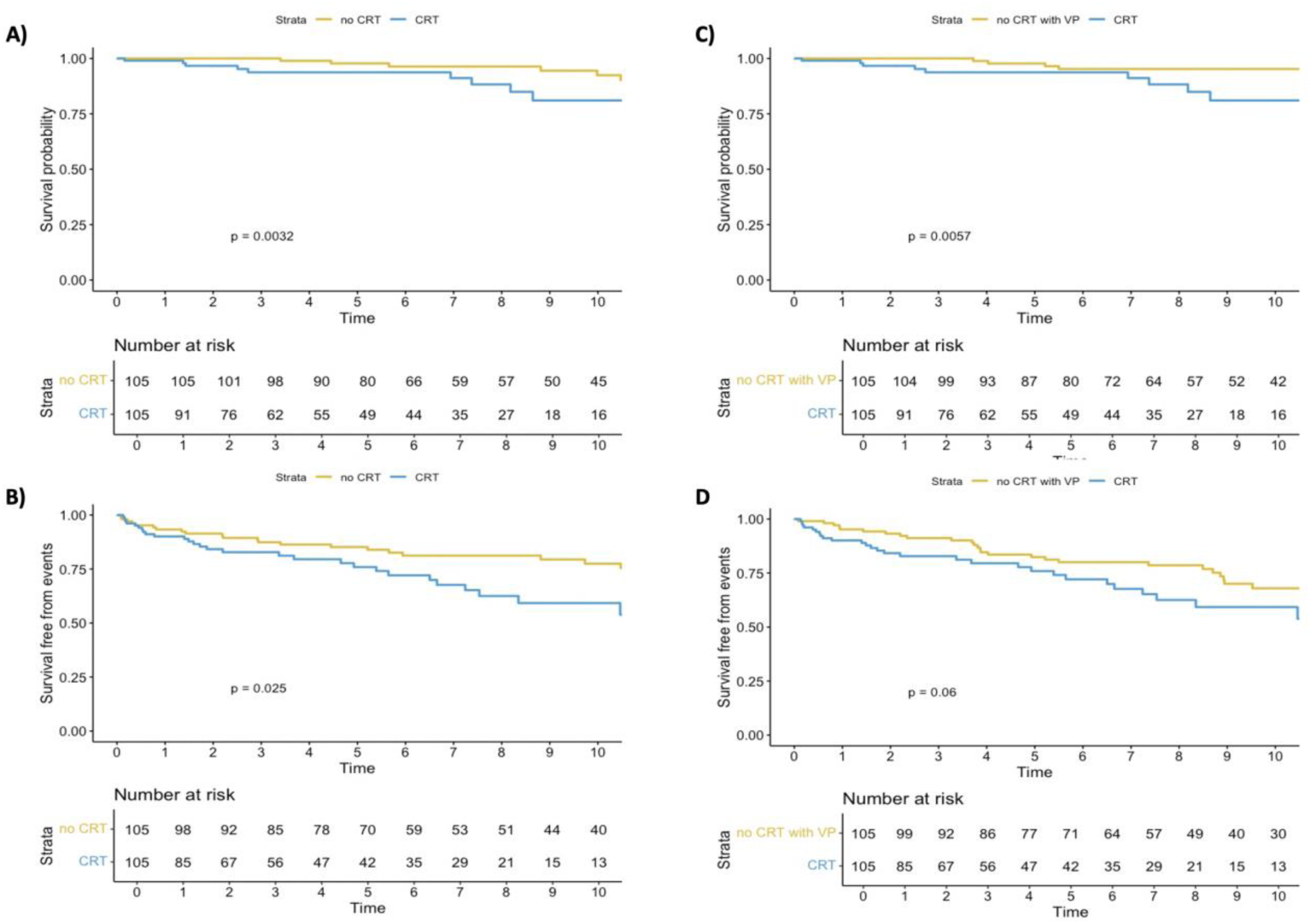
Kaplan-Meier curves for survival and survival free from events in the study population and matched controls. A) Overall survival. B) Survival free from events. C) Overall survival using only patients with ventricular pacing as matched controls. D) Survival free from events using only patients with ventricular pacing as matched controls.

**Table 5.** Events during follow-up in the study population stratified by CRT.

|  | **CRT**<br>**(N=105)** | **No CRT**<br>**(N=1616)** | **p-value** |
| --- | --- | --- | --- |
| Death | 12(11.4%) | 112(6.9%) | 0.08 |
| - Worsening HF | 9(9%) | 38(2.3%) |  |
| - Non cardiac cause | 1(1%) | 19(1.1%) |  |
| - Sudden/arrhythmic death | 0 | 23(1.4%) |  |
| - Complications from invasive procedures | 0 | 9(0.5%) |  |
| - Unknown | 2 (1%) | 0 |  |
| Hospitalization for HF | 15(14%) | 207(13%) | 0.7 |
| Sustained VT/ICD therapy | 13(12%) | 158(9.7%) | 0.4 |
| Transplant | 4(4%) | 34(2%) | 0.1 |
| Mechanical circulatory support | 3(3%) | 15(0.9%) | **0.03** |
| Composite endpoint | 29(28%) | 371(23%) | 0.2 |

Abbreviations: CRT=cardiac resynchronization therapy, HF=heart failure, ICD=implantable cardioverter-defibrillator, VT=ventricular tachycardia

**Table 6.**
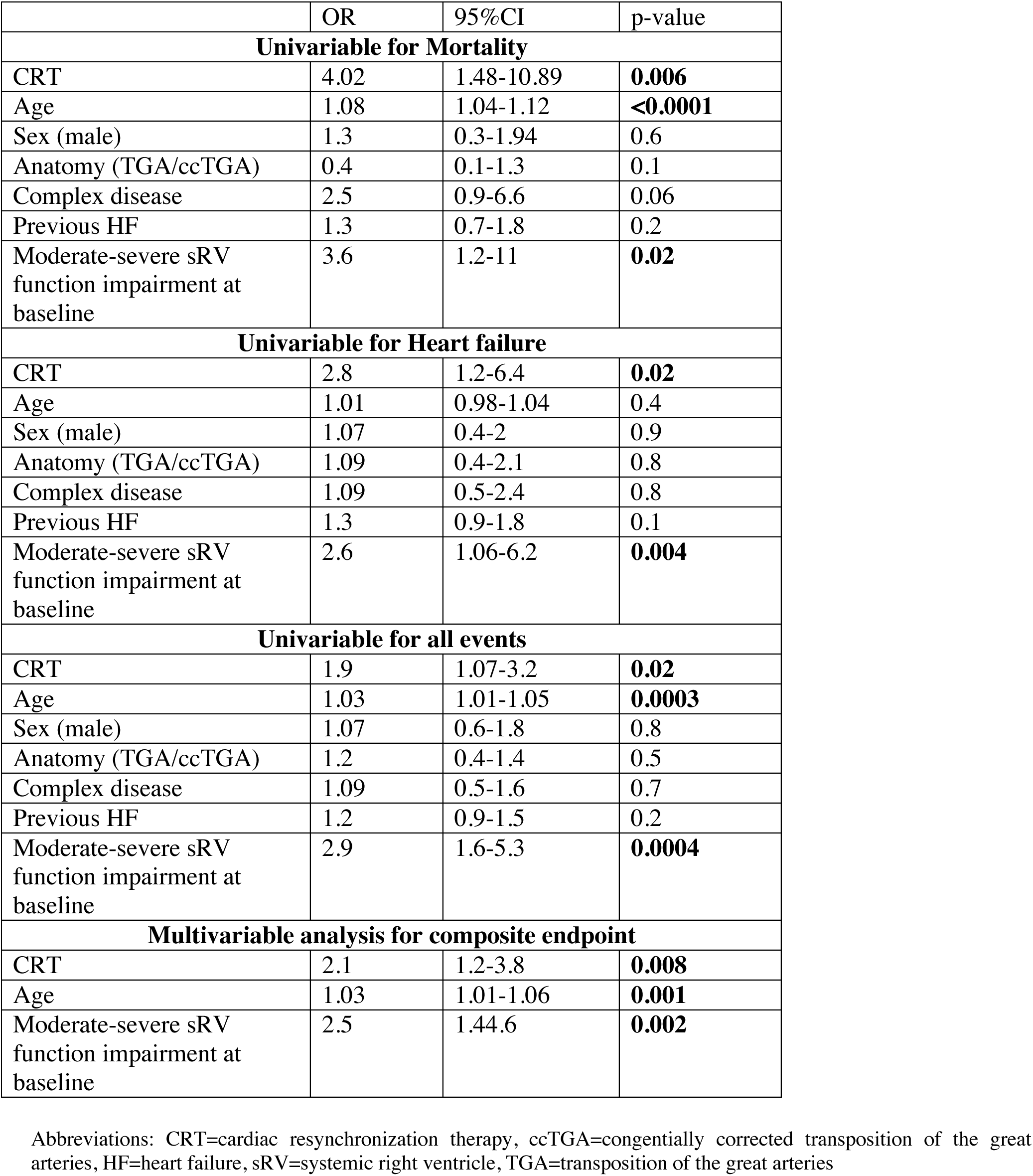
Univariable analysis for outcome events after propensity score matching.

**Table 7.** Univariable and multivariable analysis for outcome events after propensity score matching including only patients with ventricular pacing as controls.

|  | OR | 95%CI | p-value |
| --- | --- | --- | --- |
| **Univariable for Mortality** |  |  |  |
| CRT | 3.7 | 1.4-10.1 | **0.009** |
| Age | 1.05 | 1.02-1.09 | **0.0003** |
| Sex (male) | 1.05 | 0.4-2.4 | 0.9 |
| Anatomy (TGA/ccTGA) | 0.3 | 0.06-1.2 | 0.08 |
| Complex disease | 4.1 | 1.6-12.5 | **0.01** |
| Previous HF | 1.9 | 1.4-2.6 | **<0.0001** |
| Moderate-severe sRV function impairment at baseline | 1.9 | 0.7-5.3 | 0.2 |
| **Univariable for all events** |  |  |  |
| CRT | 1.6 | 0.9-2.7 | 0.06 |
| Age | 1.04 | 1.6-1.06 | **<0.0001** |
| Sex | 1.02 | 0.6-1.7 | 0.9 |
| Anatomy (TGA/ccTGA) | 0.8 | 0.5-1.4 | 0.6 |
| Complex disease | 1.1 | 0.7-1.9 | 0.6 |
| Previous HF | 1.7 | 1.3-2.1 | **<0.0001** |
| Moderate-severe sRV function impairment at baseline | 3.2 | 1.8-5.7 | **<0.0001** |
| **Multivariable for all events** |  |  |  |
| CRT | 1.9 | 1.1-3.3 | **0.02** |
| Age | 1.03 | 1.01-1.05 | **0.001** |
| Previous HF | 1.5 | 1.2-2 | **0.002** |
| Moderate-severe sRV function impairment at baseline | 2.5 | 1.3-4.4 | **0.003** |

Abbreviations: CRT=cardiac resynchronization therapy, ccTGA=congenitally corrected transposition of the great arteries, HF=heart failure, sRV=systemic right ventricle, TGA=transposition of the great arteries

### 3.5 Outcome in patients according to QRS improvement and baseline indications to CRT

Despite no overall QRS improvement, QRS duration decreased in those with previous pacing, while a QRS worsening was evident in the de-novo CRT group. A QRS narrowing>10% following CRT implantation was reported in 18 out of 86 (26%) patients with available data pre and post implant. Baseline and outcome differences between patients who did and did not show improved QRS are shown in Table 8: those with QRS improvement had longer baseline QRS duration (p=0.0001) and were more likely to have previous subpulmonary left ventricle pacing (p<0.0001). As for the outcome, no significant differences in the occurrence of events were found between the two groups. However, patients with longer QRS at baseline showed non-significantly better survival free from events after CRT implant (Supplemental Figure 3). Dividing CRT patients in two groups according to a baseline CRT indication with at least a IIa recommendation level according to the latest guidelines for pacing in CHD (8), 77(73%) were deemed fully match CRT indication considering a QRS duration ≥150 ms, sRV dysfunction, and ventricular pacing need. However, patients with CRT indication did not show lower incidence of adverse events (Table 9).

**Table 8.**
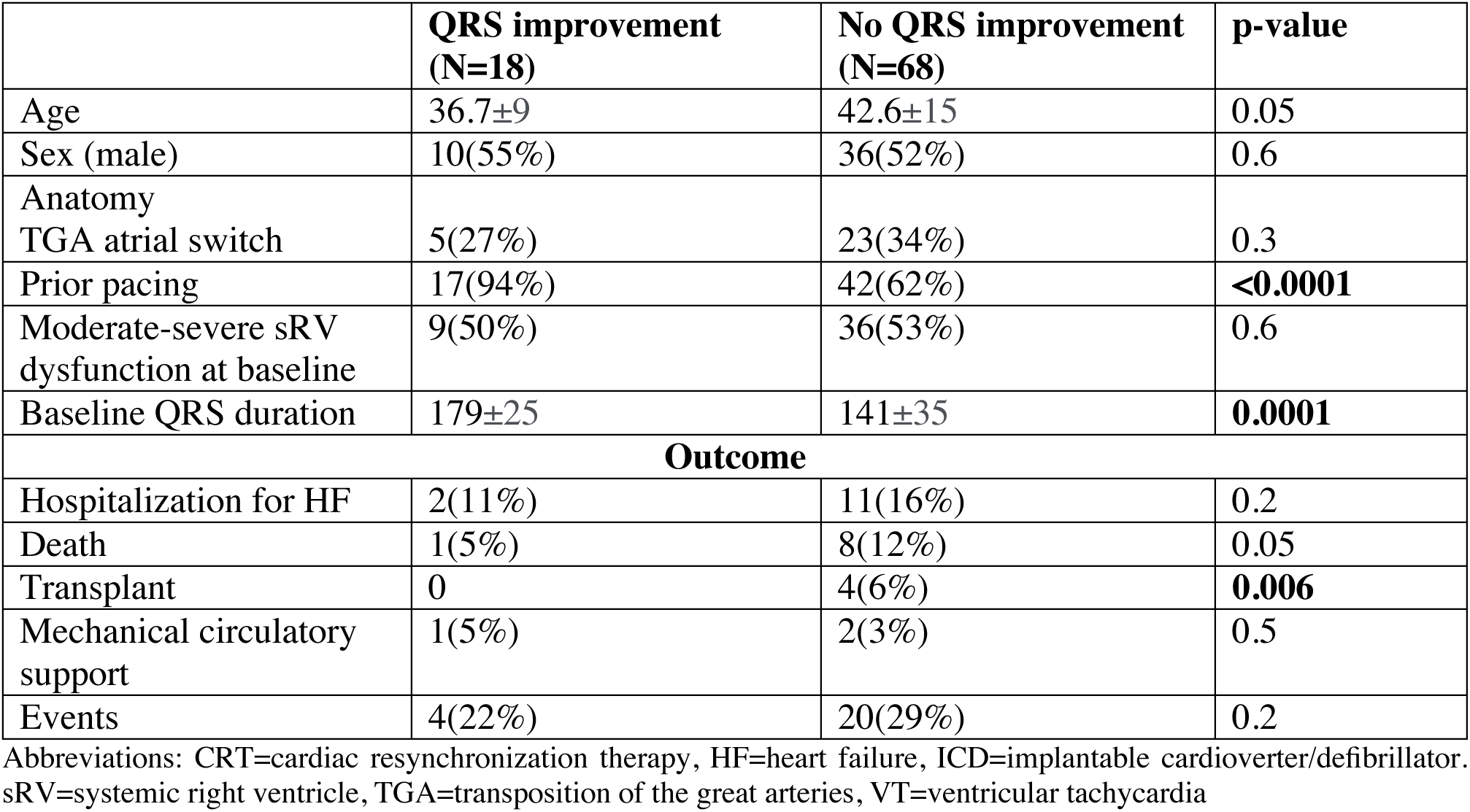
Baseline differences and outcome events in CRT patients stratified by >10% decrease from baseline QRS.

|  | **QRS improvement**<br>**(N=18)** | **No QRS improvement**<br>**(N=68)** | **p-value** |
| --- | --- | --- | --- |
| Age | 36.7±9 | 42.6±15 | 0.05 |
| Sex (male) | 10(55%) | 36(52%) | 0.6 |
| Anatomy |  |  |  |
| TGA atrial switch | 5(27%) | 23(34%) | 0.3 |
| Prior pacing | 17(94%) | 42(62%) | **<0.0001** |
| Moderate-severe sRV<br>dysfunction at baseline | 9(50%) | 36(53%) | 0.6 |
| Baseline QRS duration | 179±25 | 141±35 | **0.0001** |
| **Outcome** |  |  |  |
| Hospitalization for HF | 2(11%) | 11(16%) | 0.2 |
| Death | 1(5%) | 8(12%) | 0.05 |
| Transplant | 0 | 4(6%) | **0.006** |
| Mechanical circulatory<br>support | 1(5%) | 2(3%) | 0.5 |
| Events | 4(22%) | 20(29%) | 0.2 |

Abbreviations: CRT=cardiac resynchronization therapy, HF=heart failure, ICD=implantable cardioverter/defibrillator. sRV=systemic right ventricle, TGA=transposition of the great arteries, VT=ventricular tachycardia

**Table 9.** Baseline differences and outcome events in CRT patients stratified by baseline indication to CRT according to guidelines.

|  | **Indication (N=77)** | **No indication(N=28)** | **p-value** |
| --- | --- | --- | --- |
| Age | 44±14 | 39±11 | 0.09 |
| Sex (male) | 40(52%) | 15(54%) | 0.8 |
| Anatomy |  |  |  |
| TGA atrial switch | 23(30%) | 11(39%) | 0.4 |
| Prior pacing | 71(92%) | 0(0%) | **<0.0001** |
| Moderate-severe sRV dysfunction at baseline | 45(58%) | 7(25%) | **0.003** |
| Baseline QRS duration | 159±38 | 117±34 | **<0.0001** |
| **Outcome** |  |  |  |
| Hospitalization for HF | 10(13%) | 5(18%) | 0.5 |
| Death | 11(14%) | 1(3%) | 0.1 |
| Transplant | 2(3%) | 2(7%) | 0.2 |
| Mechanical circulatory support | 1(5%) | 2(7%) | 0.7 |
| Events | 21(27%) | 8(29%) | 0.8 |

## 4. Discussion

Our study provides international real-world long-term data in the largest population with a sRV thus far reported, enabling a large albeit retrospective comparison of patients with and without a CRT. While previous studies focused mainly on CRT response in term of EF, QRS duration or symptoms (7, 9–12), our study uses survival as an endpoint in sRV patients with and without CRT. Yet in this non-randomized cohort, worse outcomes were observed in the CRT group, as no doubt use of CRT identifies a higher risk population. Yet, despite propensity score matching accounting for baseline differences between groups that would likely be factors leading to a decision to offer CRT, and exploring many possible comparisons, we could not demonstrate any favorable improvements from CRT.

Several hypothesis could provide an explanation to our findings. Firstly, despite our best attempt at a propensity matching, the results could still be influenced by some residual confounding factors, which could not be controlled due to the retrospective nature of our study. In addition, in our sRV population, only in a minority of patients resynchronization was effectively achieved, as demonstrated by QRS duration improvement, therefore patients may likely have had residual dyssynchrony. Furthermore, in our population a non-negligible proportion of patients had a baseline QRS duration <150 ms and there is some currently emerging evidence of no beneficial effects or even potential harm below this threshold (13). The high prevalence of atrial fibrillation in our cohort may have impacted negatively on our results (14–15). Finally, consideration for CRT may be a marker of a worse disease progression trajectory and worsening HF, which may not be efficiently captured by propensity matching.

### 4.1. CRT challenges and indication in CHD

CRT has been demonstrated as an effective treatment in patients with acquired heart disease. In particular, CRT is strongly recommended for symptomatic patients with HF and EF<35% despite optimal medical therapy who have left bundle branch block and wide QRS duration (16). However, CRT use in ACHD patients is significantly limited by complex anatomy and heterogenous physiology portending unique technical challenges during implantation procedures with consequent increased complication rates and absence of randomized studies assessing CRT efficacy. Despite those limitations in the ACHD population, a recent propensity score-matched analysis on a large retrospective cohort including 167 and 324 eligible controls demonstrated a significantly improved transplant-free survival in the CRT group with significantly higher EF and shorter QRS duration at 5 year-follow-up (17). This key study supports the use of CRT in congenital patients. However, it is worth noting that the study included both pediatric and adult patients with a diagnosis of CHD or cardiomyopathy with a median age of 12 years and only 15 patients had a sRV (17).

### 4.2. CRT in sRV

Patients with a sRV pose an extra challenge for biventricular pacing due to the highly variable anatomy in this subgroup with high prevalence of associated defects. In d-TGA, access to the coronary sinus may be complicated by the presence of the atrial baffle and it typically wraps around the subpulmonary RV, and therefore an epicardial system is usually required to pace the sRV. In ccTGA, patients may frequently present with anatomical variants of cardiac veins, including coronary sinus displacement from the atrioventricular groove and/or ostial atresia (18–19). The first experience with CRT in a small group of 8 individuals with sRV from pediatric to adult age showed promising results with improved symptoms, QRS duration and sRV function suggesting potential beneficial effects from electrical therapy (9). However, following studies with larger cohorts have not consistently confirmed the initial positive results, suggesting different long-term responses to CRT in ACHD patients with sRV or systemic left ventricle in terms of systolic function (10–11). In a more recent French multicenter study including 85 patients with CHD, 36% of whom with a sRV, CRT responder rates were comparable in patients with sRV and systemic left ventricle (12). It is worth noting that the variability in response rates may be at least partially attributable to the differing definition of positive CRT response across studies. In the French study a CRT response was defined as at least 10% EF increase and/or symptoms improvement. Furthermore, EF was measured by echocardiography with Simpson’s biplane method, which has non negligible limitations in this setting for sRV geometry. Given the uncertain beneficial effects, there are no Class I indications for CRT in patients with a sRV according to a joint consensus paper dated 2014 from the Pediatric and Congenital Electrophysiology Society and the Heart Rhythm Society (HRS) (8). The sole Class IIA indication consists of a sRV EF<35% in the setting of a QRS>150 ms, RV dilation, and NYHA class II to IV symptoms (8). Kharbanda et al. (7) have previously described a multicenter retrospective experience on CRT use in a population of 80 adults with a sRV: early improvement in QRS duration and NYHA class were demonstrated exclusively in patients with previous left ventricular pacing, despite no effects on overall survival. The latter finding appears consistent with our results indicating no survival benefit in patients who underwent upgrading to CRT compared to a matched control group with left ventricular pacing. Moreover, both Kharbanda’s study and ours showed similar 5-year mortality and proportion of HF admission, supporting consistency across studies. Use of NYHA class reported by Kharbanda et al. has well-acknowledged limitations in ACHD patients (20) and was not included in our study protocol. Our data on BNP values and cardiopulmonary exercise test class improvement, although available in only a limited group of patients, provide more solid evidence of absence of disease progression following CRT implant.

### 4.3 Is CRT truly a determinant of adverse outcome in sRV?

In our cohort, CRT was a univariable predictor of death and events at follow-up and it retained significant association at multivariable analysis for our composite endpoint. However, as stated, multiple confounders likely impact the findings. Clearly, CRT was pursued in patients with indicators of more advanced heart failure, and thus was itself an independent marker of outcome. Globally, patients considered for CRT are usually in worse clinical conditions and may likely have a history of multiple HF admissions and so they are reasonably expected to have a higher incidence of end-stage HF, requiring MCS or transplant and ventricular tachycardia or death. Yet, despite matching for relevant clinical variables that may confound the results, CRT remained statistically associated with 2-3-fold higher risk for the combined outcome in our study cohort. Perhaps this is not surprising given that the average QRS duration worsened, the opposite desired effect from CRT. Regardless, despite the retrospective nature of the study, the data suggest that the appropriate selection of potential CRT candidates with sRV remains to be elucidated.

Moreover, despite no QRS improvement in the long-term, BNP values, peakVO2, and tricuspid regurgitation severity, when available, remained largely stable at follow-up, supporting the hypothesis that CRT may attenuate disease progression in some. In agreement with previous studies, no differences in outcome were demonstrated in CRT patients with d-TGA or ccTGA anatomy or epicardial/transvenous leads (7). Additionally, despite previous studies suggesting a greater benefit from CRT in patients upgraded from subpulmonary ventricle pacing (7), in our population CRT patients matched solely with subpulmonary ventricle paced patients had worse survival.

Several points may be considered potentially explanatory of the paradoxical increased incidence of events among CRT patients. First, individuals with a failing sRV may be referred too late for CRT, as suggested in our cohort by the severe delay from pacemaker implant to CRT upgrading with 1096 person-years spent with subpulmonary ventricle pacing. Second, patients’ selection pathway for CRT may be not appropriate for those with sRV physiology. Third, the different electrical activation of a sRV may interfere with biventricular pacing efficacy. In particular, CRT was initially evaluated and recommended mainly for HF treatment in patients with left bundle branch block, while most sRV patients have a right bundle branch block. As for the delayed consideration for CRT in sRV, this is likely a global issue evidenced also by previous studies with cohorts already in advanced HF at time of implant (7).

### 4.4 Future directions

Recent data from conduction system pacing (CSP) showed promising results. The largest study to date including 65 ACHD patients, though none of them having a sRV, demonstrated similar results from CSP and conventional CRT in terms of EF improvement with greater QRS narrowing in the CPS group (21). Previous studies in smaller groups had already showed feasibility with good immediate results and low complications rate from CSP in patients with a sRV (22–25). According to this evidence, recent HRS guidelines on cardiac physiologic pacing for the avoidance and mitigation of heart failure support CRT implant in patients with CHD and sRV with HF symptoms and CSP in those with ccTGA requiring pacing for advanced atrioventricular block (26). Future studies may ascertain whether improved patient selection for CRT or use of CSP, which may represent a more “physiological” cardiac stimulation, may be more effective in improving the outcome of patients with a failing sRV.

### 4.5. Limitations

There are several limitations to the present analysis. Due to the retrospective nature of the study, this analysis is subject to variable data quality and degree of missingness related to data entry at participating sites, as well as to medical technology and knowledge from a prior era. Residual confounding from factors that were unmeasured or not included in the matching process may be present. In addition, the decision pathway for CRT referral may vary greatly across the participating centres, and therefore our results may have been influenced by potential indication bias, which is notoriously difficult to eliminate from retrospective studies. CRT patients may have been captured at a later stage compared to controls. Furthermore, baseline data were not necessarily obtained concurrently with the timing of CRT implant, and may not have reflected the complete clinical picture at the time. Echo-derived sRV function was assessed using a semiquantitative designation (normal, mild, moderate, or severe), in order to overcome differences between centres. CMR was not consistently performed, especially in those with these devices. Another limitation is that patients could not be matched for some variables (BNP values, peakVO2, tricuspid regurgitation severity) due to missing data. Propensity score matching focused on variables with more universal acquisition, increasing the potential for residual confounding. Despite those limitations, the finding of a notable association with worse, rather than better, outcomes suggest that CRT is likely to be of little or no benefit as it has been applied in clinical practice among the sRV population. There remains the possibility, however, that particular approaches (e.g., specific timing or type of intervention, and specific patient sub-populations) may nevertheless provide benefit.

## 5. Conclusions

In this international, retrospective study of a large cohort with a sRV, CRT implantation was not associated with improvement in overall survival nor with reduced incidence of cardiovascular events, even after propensity-matching. Few showed actual improvement in QRS duration, consistent with this lack of effect. Incidence of complications was not negligible, though most were mild. Further studies are required to better define candidates with a sRV most likely to derive benefits from CRT, explore alternative CRT strategies, and elucidate the optimal timing of this intervention.

## Data Availability

Data will be available from corresponding author upon reasonable request

---

**Supplemental Figure 1.**
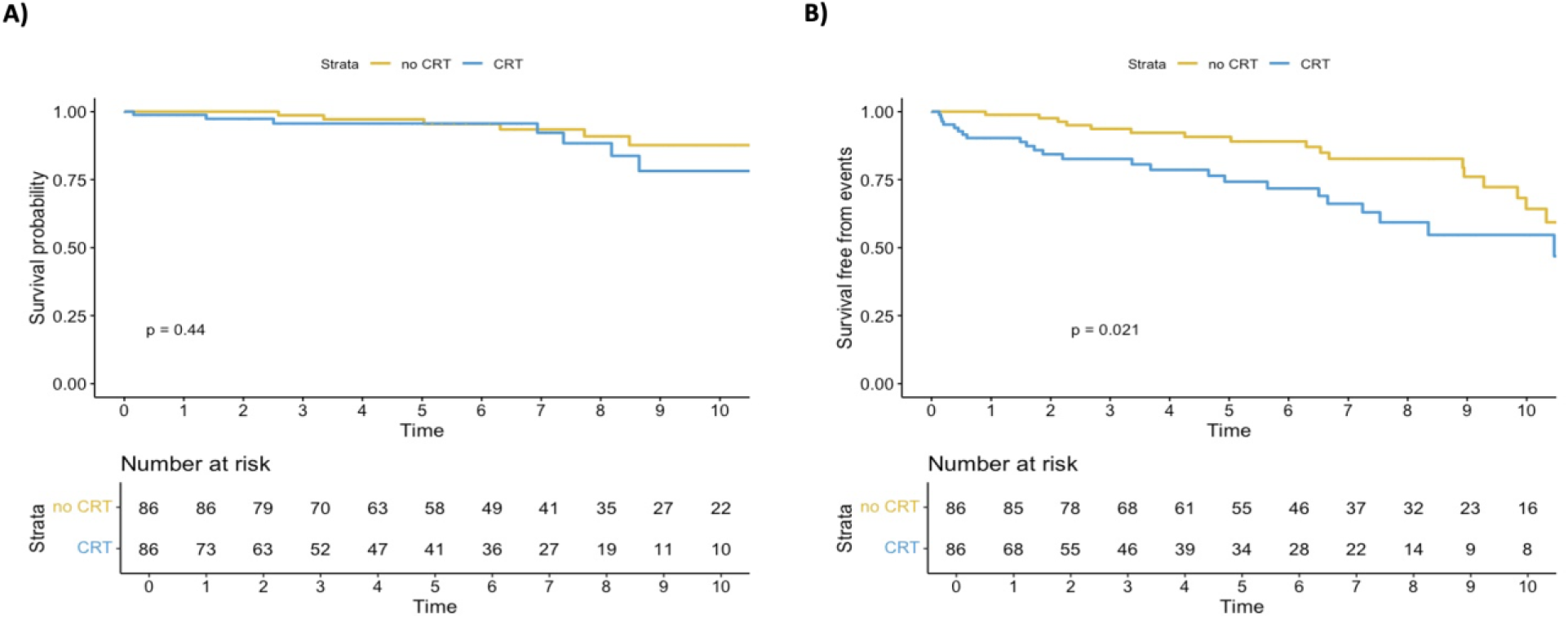
Kaplan-Meier curves for survival and survival free from events in CRT patients matched with controls for age, sex, anatomy (ccTGA/TGA), complex disease, previous admission for heart failure, sRV systolic function and QRS duration at baseline.

**Supplemental Figure 2.**
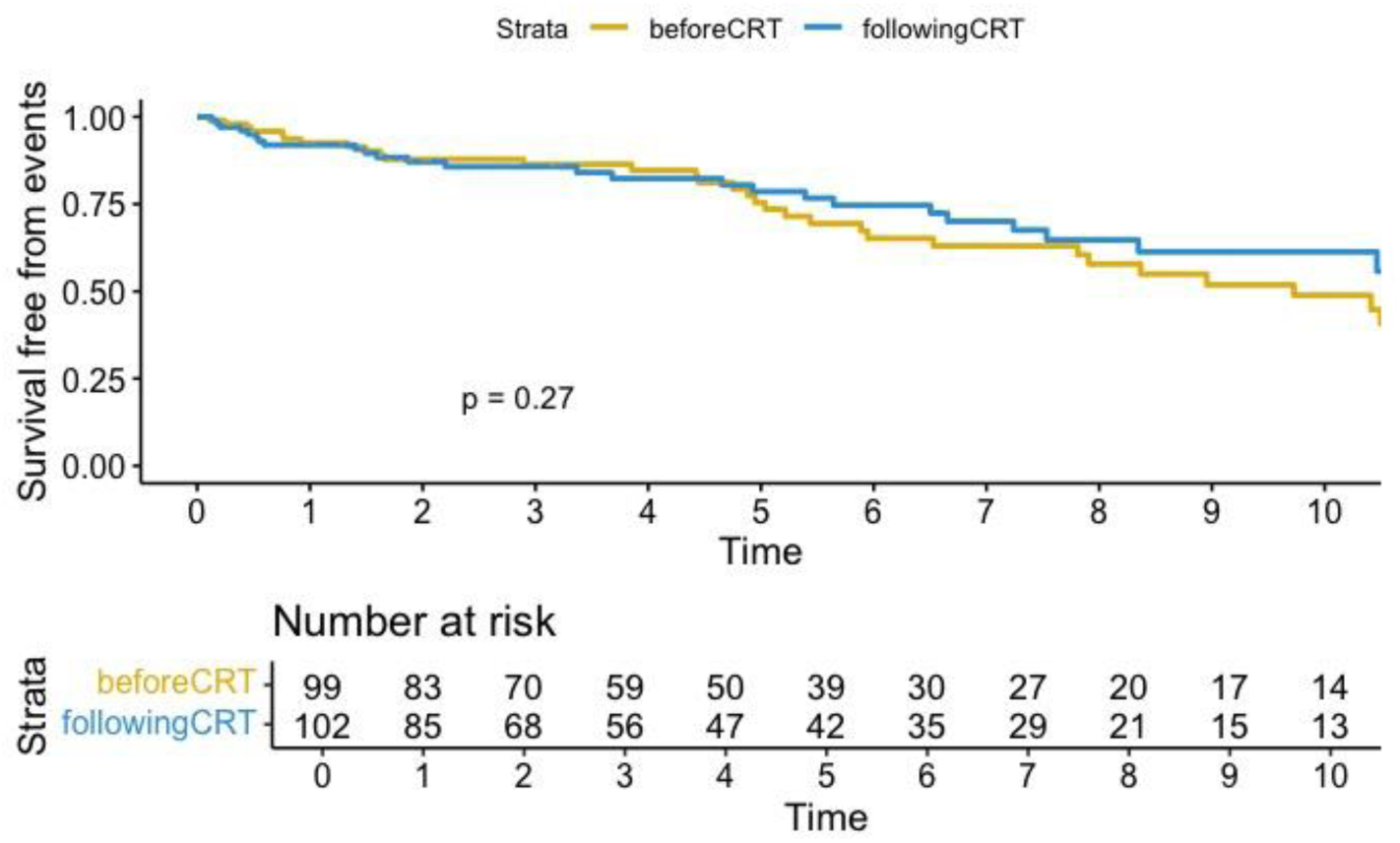
Kaplan-Meier curves survival free from events in CRT patients before and after implant.

**Supplemental Figure 3.**
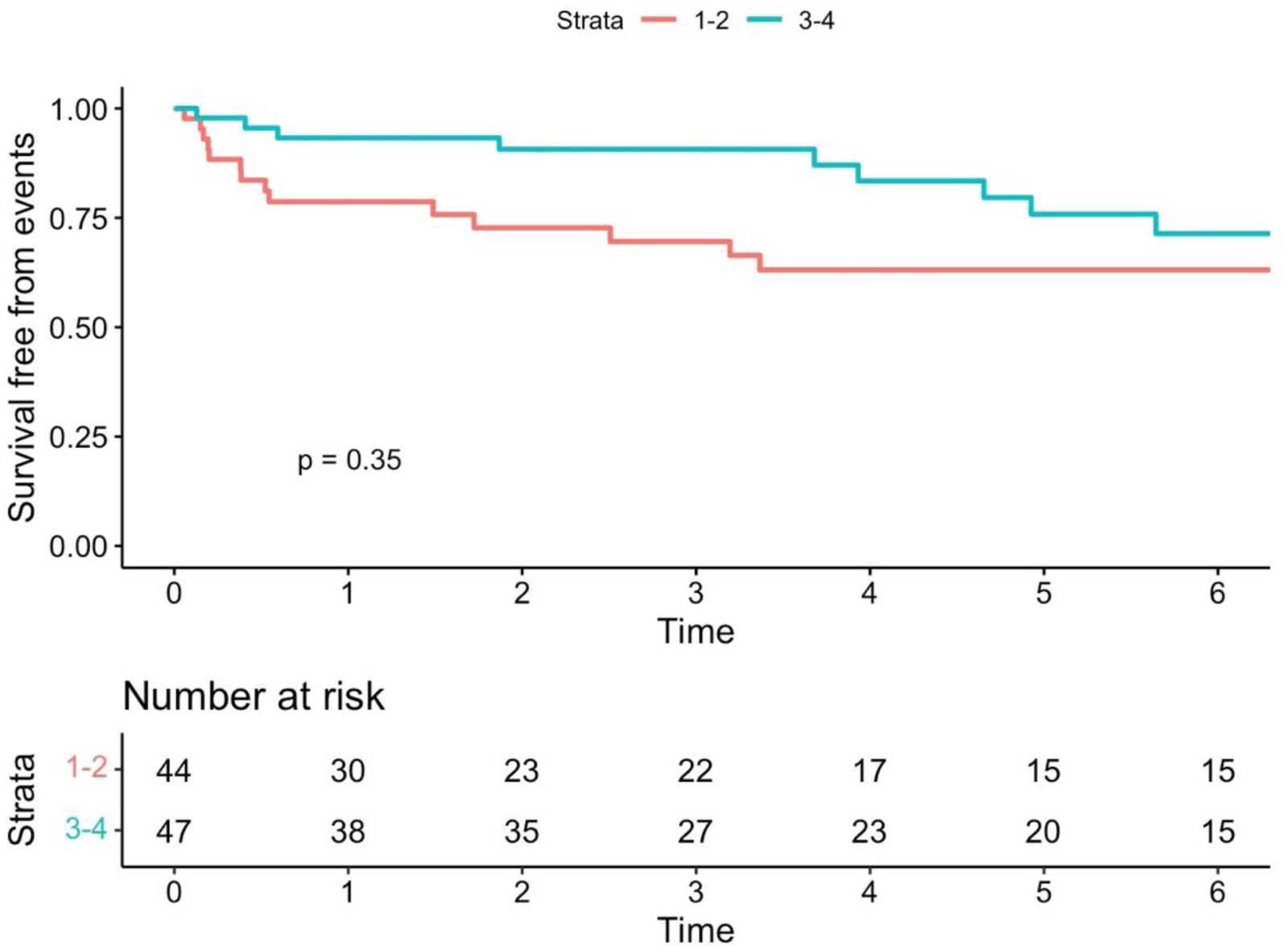
Kaplan-Meier curve for survival free from events in CRT patients stratified by baseline QRS duration. 1-2= first and second quartile; 3-4=third and fourth quartile

**Supplemental Table 1.** Events occurrence in de-novo CRT compared to CRT upgraded patients and in patients with transvenous compared to epicardial CRT.

|  | **Patients with prior pacing (N=71)** | **De novo CRT (N=34)** | **p-value** |
| --- | --- | --- | --- |
| Hospitalization for HF | 19(27%) | 12(35%) | 0.4 |
| Sustained VT/ICD therapy | 22(31%) | 12(35%) | 0.6 |
| Death | 7(10%) | 4(12%) | 0.7 |
| Transplant | 2(2%) | 2(6%) | 0.2 |
| Mechanical circulatory support | 1(1.4%) | 2(6%) | 0.1 |
| Events | 36(50%) | 20(58%) | 0.4 |
|  | **Patients with transvenous lead (N=36)** | **Patients with epicardial lead (N=17)** | **p-value** |
| Hospitalization for HF | 12(33%) | 5(29%) | 0.7 |
| Sustained ventricular tachycardia/ICD therapy | 8(22%) | 6(33%) | 0.4 |
| Death | 3(8%) | 1(6%) | 0.7 |
| Transplant | 2(6%) | 0 | 0.3 |
| Mechanical circulatory support | 2(6%) | 1(6%) | 1 |
| Events | 16(44%) | 9(53%) | 0.5 |

Abbreviations: CRT=cardiac resynchronization therapy, HF=heart failure, ICD=implantable cardioverter-defibrillator, VT=ventricular tachycardia

**Supplemental Table 2.** Baseline differences and outcome events in CRT patients stratified by baseline anatomy.

|  | **TGA (N=34)** | **ccTGA (N=71)** | **p-value** |
| --- | --- | --- | --- |
| Age | 37±10 | 39.9±15 | 0.3 |
| Sex (male) | 17(50%) | 38(53%) | 0.6 |
| Anatomy |  |  |  |
| TGA atrial switch | 17(39%) | 25(43%) | 0.7 |
| Prior pacing | 22(65%) | 49(69%) | 0.6 |
| Moderate-severe sRV dysfunction at baseline | 16(47%) | 36(51%) | 0.7 |
| **Outcome** |  |  |  |
| Hospitalization for HF | 5(15%) | 10(14%) | 0.9 |
| Death | 2(6%) | 10(14%) | 0.2 |
| Transplant | 2(6%) | 2(3%) | 0.4 |
| Mechanical circulatory support | 0 | 3(4%) | 0.2 |
| Events | 9(26%) | 20(28%) | 0.8 |

Abbreviations: CRT=cardiac resynchronization therapy, HF=heart failure, ICD=implantable cardioverter/defibrillator. sRV=systemic right ventricle, TGA=transposition of the great arteries

